# Evidence for the utility of cfDNA plasma concentrations to predict disease severity in COVID-19

**DOI:** 10.1101/2021.04.29.21256291

**Authors:** Katharina Hoeter, Elmo Neuberger, Susanne Fischer, Manuel Herbst, Ema Juškevičiūtė, Heidi Rossmann, Martin F. Sprinzl, Perikles Simon, Marc Bodenstein, Michael K.E. Schäfer

## Abstract

COVID-19 is a pandemic caused by the highly infective SARS-CoV-2. There is a need for biomarkers not only for overall prognosis but also for predicting the response to treatments and thus for improvements in the clinical management of patients with COVID-19. Circulating cell-free DNA (cfDNA) has emerged as a promising biomarker in the assessment of various disease conditions. The aim of this retrospective and observational pilot study was to examine the potential value of cfDNA plasma concentrations as a correlative biomarker in hospitalized COVID-19 patients. Lithium-Heparin plasma samples were obtained from twenty-one COVID-19 patients during hospitalization in the University Medical Center of Mainz, Germany, and the cfDNA concentrations were determined by quantitative PCR yielding amplicons of long interspersed nuclear elements (LINE-1). cfDNA plasma concentrations of COVID-19 patients ranged between 247.5 and 6346.25 ng/ml and the mean concentrations were 1831 ± 1388 ng/ml (± standard deviation). Correlations were found between cfDNA levels and the occurrence of acute respiratory distress symptom (ARDS), acute kidney injury (AKI), myositis, neurological complications, bacterial superinfection and disease severity as defined by sepsis-related organ failure assessment score (SOFA) score. D-Dimer and C-reactive-protein (CRP), determined by clinical laboratory analysis, showed the highest correlations with cfDNA levels. The results of this observational study suggest that cfDNA plasma concentrations may serve as a predictive biomarker of disease severity in COVID-19. Prospective studies enrolling larger patient cohorts are ongoing to test this hypothesis.

## Introduction

Coronavirus disease 2019 (COVID-19) is a pandemic caused by the highly infective SARS-CoV-2. To date, SARS-CoV-2 has caused more than 130 million cases worldwide. Although COVID-19 vaccines are being developed rapidly, compared to traditional vaccines, and have been approved worldwide (1), the ongoing outbreak of COVID-19 puts an enormous strain on health resources and represents an extraordinary threat to global public health (2). In addition, new SARS-CoV-2 variants with increased transmission rate have emerged in the past months which further complicated the situation (3). Disease caused by SARS-CoV-2 infection ranges from mild course of illness to severe respiratory diseases, multiple organ failure and death. Amongst others, pulmonary manifestations are common and range from cough to pneumonia and acute lung failure. Hematological and immune system-related changes such as thrombocytopenia and dysregulation of blood clotting have been reported (4). In addition, neurological manifestations (5), acute kidney failure (6) and symptoms of the gastrointestinal tract such as nausea and vomiting, diarrhea, and gastrointestinal bleeding occur in the context of COVID-19 disease (7). Encouraging effects on the course of the disease have been reported after corticosteroid treatment (8), however, to date, there are no generally proven effective therapies for COVID-19 or antivirals against SARS-CoV-2 (2). Thus, the only life-saving therapy is bridging-to-recovery which means in the case of organ failure by organ support or replacement.

The increasing incidence of SARS-CoV-2-infections with possible serious consequences in almost every age group, makes optimal biomarkers necessary. Biomarkers are not only necessary for prognosis, but also for predicting the response to treatments and thus for improvements in the clinical management of patients with COVID-19. Recent studies associated laboratory measures of hyperinflammation such as macrophage chemoattractant protein 1 (MCP-1), C-reactive-protein (CRP), and interleukin-6 (IL-6) as strong predictors of disease severity in hospitalized patients with COVID-19 (9, 10). Another promising, non-invasive biomarker from liquid biopsy is cell-free DNA (cfDNA), which is passively released after cell damage and/or actively released from hematopoietic (immune) cells (11, 12). Elevated concentrations of cfDNA have been detected under various pathological conditions (13). In tumour diseases, cfDNA levels have been utilized to evaluate tumour burden, progression, and treatment responses (14-17). Elevated levels of cfDNA were also found in patients with severe bacterial infections or viral infections and correlated with the course and severity of diseases, respectively (18, 19). Most notably, recent studies (20, 21) report on the value of cfDNA as a predictive biomarker for COVID-19 severity. Here, we present data from a retrospective observational study to further assess the value of cfDNA as a potential biomarker in hospitalized COVID-19 patients.

## Methods

### Patients

Twenty-one patients hospitalized between March and June 2020 at the University Medical Center, Mainz, Germany, were assessed in this explorative study. Patients` clinical data and laboratory findings were reviewed retrospectively through the electronic hospital information systems (i.s.h.med^®^, SAP, Weinheim Germany, Nexus Swisslab, Berlin, Germany). The study was approved by German law [Landeskrankenhausgesetz §36 and §37] in accordance with the Declaration of Helsinki and by the local Ethics Committee of “Landesärztekammer Rheinland-Pfalz” (reference numbers 2020-15116-retrospective).

SARS-CoV-2 infection was confirmed by polymerase chain reaction (PCR) from respiratory samples and all other laboratory assays were performed in the accredited (DIN-ISO 15.189) Institute of Clinical Chemistry and Laboratory Medicine of the University Medical Center, Mainz, as described (22).

### Blood sample processing and quantification of cfDNA

Lithium-Heparin syringes (S-Monovette, Sarstedt) were used for collection of blood plasma samples and centrifuged within 2-3 hours after collection at 3746 x g for 10 min at room temperature. Plasma aliquots were stored at −80°C prior to quantification of cfDNA concentrations. The plasma cfDNA concentrations were determined according to Neuberger et al. (preprint, medRxiv, https://doi.org/10.1101/2021.01.17.21249972 using quantitative real-time PCR (qPCR). The qPCR assay targets a 90bp hominoid specific repetitive element of the long interspersed element 1 (LINE1) family 2 (L1PA2). The assay shows a limit of quantification < 1ng/ml, a sufficient dynamic range, as well as repeatability < 12%. To determine storage dependent changes in cfDNA levels in lithium heparin syringes, EDTA or Lithium-Heparin syringes containing plasma from three different subjects were collected and processed directly, or stored for five days at 4°C, respectively. As expected, in EDTA samples the cfDNA concentration increased significantly ∼60 fold during the extended storage time (P=0.004). In Lithium-Heparin samples no significant differences were determined between direct processing and extended storage (P=0.5). The samples showed a concentration of 71.8 ± 33.1 and 80.2 ± 32.1 ng/ml, respectively (mean ± SD) demonstrating that Lithium-Heparin rather than EDTA plasma samples are suitable for the analysis of cfDNA.

### Statistical analysis

Data analysation is descriptive. Mean and standard deviation or absolute and relative frequency were calculated. Student`s T-test was used to describe association between clinical complication of patients and the concentration of cfDNA in plasma samples. Spearman’s correlation was used to describe association between cfDNA concentration (grouped as low, moderate, or high) and laboratory parameters.

## Results

Twenty-one hospitalized patients with positive proof of SARS-CoV-2 were assessed in this study, twelve of whom were male and nine were female. The patients were 68 ± 17 years and on average overweight (BMI 28.8 ± 6.6 kg/m^2^, table 1).

**Table 1.** Biometric data

|  |  |
| --- | --- |
| Age (years) | 68 ± 17 |
| Sex | 12 male. 9 female |
| Height (cm) | 174.1 ± 8.2 |
| Weight (kg) | 87.5 ± 22.8 |
| BMI (kg/m<sup>2</sup>) | 28.8 ± 6.6 |

Data are given as mean ± standard deviation. *BMI* body mass index

Most of the patients had at least one pre-existing risk-factor for a severe course of COVID-19 comprising cardiac (19%), renal (33%), pulmonary (38%), or immunological conditions (14%), arterial hypertension (57%), diabetes (19%) or adipositas (33%, table 2).

**Table 2.** Pre-existing condition

| Type of condition | Frequency [n(%)]<br>(*n*=21) | Distribution of sexe<br>[f/m] |
| --- | --- | --- |
| Cardiac | 4 (19%) | 1/3 |
| Renal | 7 (33%) | 3/4 |
| Pulmonary | 8 (38%) | 3/5 |
| Immunological | 3 (14%) | 1/2 |
| Arterial Hypertension | 12 (57%) | 3/9 |
| Diabetes mellitus Typ II | 4 (19%) | 2/2 |
| Adipositas (BMI $\geq 30$ kg/m<sup>2</sup>) | 7 (33%) | 4/3 |

*BMI* body mass index. *f* female. *m* male

Several laboratory parameters of the patients were elevated during their course of COVID-19 (table 3). Increased levels of creatinine, urea or LDH and decreased estimated glomerular filtration rate (eGFR) were indicative for organ dysfunction, i.e. acute kidney injury (AKI). Inflammatory parameters CRP and procalcitonin (PCT) were markedly present in the majority of patients, reflecting hyperinflammation and/or secondary bacterial infection. Increased D-Dimer concentration was considered as an indicator for COVID-19 associated coagulopathy (table 3).

**Table 3.** Laboratory parameters reflecting organ dysfunction (creatinine, urea, LDH), inflammation (CRP, PCT) or COVID-19 associated coagulopathy (D-Dimer)

| Laboratory parameter | Mean ± SD | [Min. – Max.] | Normal range |
| --- | --- | --- | --- |
| Maximum D-Dimer (mg/l) | 7.59 ± 8.09 | [0.55 – 28.69] | < 0.50 |
| Maximum creatinine (mg/dl) | 2.49 ± 2.23 | [0.57 – 7.24] | 0.73-1.18 |
| Maximum urea (mg/dl) | 50.71 ± 34.28 | [6 – 106] | 9-21 |
| Lowest eGFR (ml/min/m) | 50.95 ± 36.29 | [7 – 120] | 68-108 |
| Maximum CRP (mg/l) | 235.7 ± 158 | [46 – 568] | <5 |
| Maximum PCT (ng/ml) | 11.56 ± 43.99 | [0.02 – 208] | <0.5 |
| Maximum LDH (U/l) | 669.67 ± 374.5 | [235 – 1849] | <245 |

*SD* standard deviation, *eGFR* estimated glomerular filtration rate, *CRP* C-reactive protein, *PCT* procalcitonin, *LDH* lactate dehydrogenase

The patients suffered from several complications, most frequently AKI (62%) followed by pulmonary complications including invasive ventilation and ARDS (43%), anemia and secondary infections (table 4). COVID-19 was potentially fatal, 2 of 21 patients died during the hospital stay (table 5).

**Table 4.** Complications

| Type of complication | Frequency [n(%)]<br>(n=21) | Distribution of sexe<br>[f/m] |
| --- | --- | --- |
| ***Pulmonary complications*** |  |  |
| ARDS | 9 (43%) | 2/7 |
| ECMO therapy | 0 (0%) | n.a. |
| Invasive ventilation ≥ 7d | 9 (43%) | 2/7 |
| Reintubation | 2 (11%) | 1/1 |
| Tracheostomy | 6 (34%) | 2/4 |
| Lowest Oxygenation-index (mmHg) | 98.33 ± 37.65 | n.a. |
| Need for invasive ventilation (days) | 20.4 ± 15.51 | n.a. |
| ***Thromboembolism*** |  |  |
| Mesenterial ischemia | 1 (5%) | 0/1 |
| Catheter associated thrombosis | 2 (10%) | 0/2 |
| Pulmonary artery embolism | 1 (5%) | 1/0 |
| ***Neurological complications*** |  |  |
| Critical illness polyneuropathy | 1 (5%) | 0/1 |
| Delirium | 7 (33%) | 2/5 |
| Relapse of pre-existing condition | 3 (14%) | 1/2 |
| Delayed wake up | 3 (14%) | 0/3 |
| ICU acquired weakness | 1 (5%) | 1/2 |
| ***Renal complications*** |  |  |
| AKI 1 -3 (KDIGO or RRT) | 13 (62%) | 3/10 |
| ***Cardiac complications*** |  |  |
| CPR | 1 (5%) | 1/0 |
| Myocardial injury | 1 (5%) | 0/1 |
| Atrial fibrillation | 2 (10%) | 0/2 |
| Angina pectoris | 1 (5%) | 0/1 |
| ***Other complications*** |  |  |
| Secondary infection | 12 (57%) | 3/9 |
| Anemia | 18 (86%) | 6/12 |
| Diarrhea | 2 (10%) | 1/1 |

*SD* Standard deviation, *AKI* acute kidney injury *KDIGO* Kidney Disease: Improving Global Outcomes,
*RRT* Renal replacement therapy, *ICU* Intensive Care Unit, *ARDS* acute respiratory distress syndrome,
*ECMO* Extra corporal membrane oxygenation, *f* female, *m* male

**Table 5.** Course of the disease

| Parameter | n(%) or mean ± SD | Distribution of sexe<br>[f/m] |
| --- | --- | --- |
| LOS ICU (d) | 28.78 ± 19.31 | |
| Readmission on ICU | 1 (5%) | 1/0 |
| LOS in-hospital (d) | 29.94 ± 19.45 | |
| Mortality [*n*(%)] | 2 (10%) | 0/2 |

*LOS* Length of Stay *ICU* Intensive Care Unit *d* days, *SD* standard deviation, *f* female, *m* male

Association between plasma cfDNA concentration and complications, risk factors or outcome revealed that the occurrence of myositis was associated with higher cfDNA concentration (table 6). Interestingly, thromboembolism was found in patients with relatively low cfDNA concentrations and higher cfDNA concentrations were associated with a lack of thromboembolism (table 6).

**Table 6.** Association between cfDNA concentration and complications, risk factors or outcome.

| Complication, risk factor or outcome | cfDNA (mean ± SD) | Significance level |
| --- | --- | --- |
| **Complications** |  |  |
| ARDS | 2010 ± 1068. n=11 | P = 0.566 |
| No ARDS | 1620 ± 1829. n=10 |  |
| Thromboembolism | 1054 ± 689. n=4 | P = 0.087 |
| No Thromboembolism | 2005 ± 1540. n=17 |  |
| AKI | 1679 ± 1998. n=13 | P = 0.767 |
| No AKI | 1914 ± 1084. n=8 |  |
| Vasopressor | 1769 ± 889. n=9 | P = 0.874 |
| No Vasopressor | 1866 ± 1808. n=12 |  |
| Delirium | 1741 ± 1069. n=10 | P = 0.806 |
| No Delirium | 1900 ± 1786. n=11 |  |
| Myositis | 2260 ± 1582. n=14 | P = 0.013 |
| No Myositis | 953 ± 553. n=7 |  |
| SOFA ≤9 | 1237 ± 596. n=3 | P = 0.266 |
| SOFA > 9 | 1853 ± 978. n=7 |  |
| **Risk factors** |  |  |
| Arterial Hypertension | 1744 ± 1124. n=12 | P = 0.794 |
| No Arterial Hypertension | 1932 ± 1879. n=9 |  |
| Diabetes mellitus | 1687 ± 1183. n=4 | P = 0.818 |
| No Diabetes mellitus | 1856 ± 1542. n=17 |  |
| BMI ≥ 30 | 1724 ± 1075. n=7 | P = 0.806 |
| BMI < 30 | 1874 ± 1648. n=14 |  |
| **Outcome** |  |  |
| Survived | 1470 ± 860. n=20 | P = 0.172 |
| Died | 5245 ± 1557. n=2 |  |

Two-sided T-test (unequal variance, Lavene), *P* significance level
*BMI* body mass index, *ARDS* acute respiratory distress syndrome, *AKI* acute kidney injury,
*SOFA* sequential organ failure assessment

The average cfDNA plasma concentration was 1831 ± 1388 ng/ml (table 7). The lowest cfDNA concentration was 248 ng/ml, the highest cfDNA concentration was 6346 ng/ml. The patients were divided into three groups according to their plasma cfDNA concentrations: High concentration > 2000 ng/ml, moderate concentration 1000-2000 ng/ml and low concentration ≤ 1000 ng/ml. The group sizes (each consisting of n=6-8) were almost evenly distributed (table 7).

**Table 7.** Groups of cfDNA concentration

| Cohort | sample size n (%) | Distribution of sexe<br>[f/m] | Mean ± SD (ng/ml) |
| --- | --- | --- | --- |
| All patients | 21 (100%) | 9/12 | 1813.1 ± 1387.96 |
| cfDNA.high<br>> 2000 ng/ml | 8 (36%) | 4/4 | 3190.1 ± 1350 |
| cfDNA.moderate<br>1000-2000 ng/ml | 7 (32%) | 1/6 | 1475.6 ± 347.5 |
| cfDNA.low<br>< 1000 ng/ml | 7 (32%) | 4/3 | 576.9 ± 204.2 |

cfDNA cell-free Desoxyribonucleic acid, SD standard deviation, *f* female, *m* male

Associations between these three groups, complications and outcome included predominantly renal, pulmonary, and neurological complications, among others, and respiratory complications were more frequently in the cohort with moderate and high cfDNA concentrations. Mortality only occurred in the group with highest cfDNA concentration (table 8).

**Table 8.**
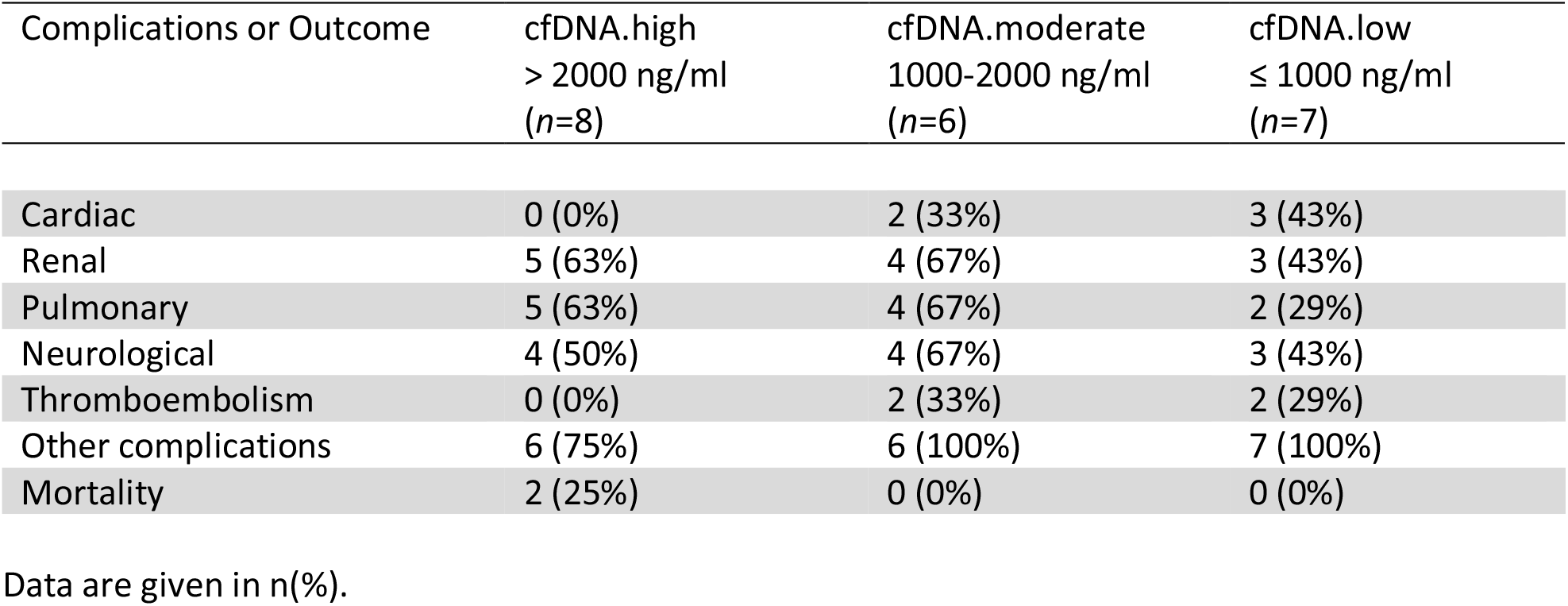
Complications and outcome in different cfDNA groups

Extreme laboratory parameters occurred in temporal association with the cfDNA concentration measurement depending on the group (table 9). D-Dimer and CRP concentration were associated to cfDNA concentration (both p-value of Spearman’s correlation were 0.08, table 9).

**Table 9.** Association between cfDNA concentration and laboratory parameters.

| Laboratory parameters<br>[unit, normal range,<br>reported value] | Low cfDNA<br>(n=7, mean) | Moderate cfDNA<br>(n=6, mean) | High cfDNA<br>(n=8, mean) | Spearman's rho | P |
| --- | --- | --- | --- | --- | --- |
| **Creatinine**<br>[mg/dl, 0.73-1.18,<br>max in 7 days] | 0.68 | 1.77 | 1.85 | 0.28 | 0.21 |
| **Bilirubin**<br>[mg/dl, 0.2-1.2,<br>max in 7 days] | 0.70 | 1.35 | 0.75 | 0.24 | 0.29 |
| **Lactate**<br>[mmol/l, 0.5-1.6,<br>max in 2 days] | 1.10 | 1.40 | 1.90 | 0.36 | 0.23 |
| **LDH**<br>[U/l, <245,<br>max in 2 days] | 386.00 | 367.50 | 481.50 | 0.38 | 0.18 |
| **D-Dimer**<br>[mg/l, <0.5,<br>max in 2 days] | 1.12 | 4.01 | 4.27 | 0.67 | 0.08 |
| **Thrombocytes**<br>[/nl, 150-360,<br>min in 2 days] | 227.00 | 263.50 | 230.00 | -0.01 | 0.98 |
| **CRP**<br>[mg/l, <5,<br>max in 2 days] | 54.00 | 65.50 | 89.00 | 0.39 | 0.08 |
| **Leucocytes**<br>[/nl, 3.5-10,<br>max in 2 days] | 8.30 | 8.67 | 8.54 | 0.10 | 0.66 |

*LDH* lactate dehydrogenase, *CRP* C-reactive protein
Spearman's rank correlation coefficient was calculated analyzing the laboratory parameters in three different cohorts of cfDNA concentration (low, moderate, high). The laboratory parameters evaluated here are maximum (max) or minimum (min) values during the next 2 or 7 days after cfDNA concentration was measured.

## Discussion

This retrospective observational study explored correlations between cfDNA plasma levels and clinical complications as well as clinical blood laboratory measures. In our patient cohort cfDNA concentration was associated to mortality, myositis, respiratory complications, D-Dimer and CRP, thus reflecting outcome, organ-complications and inflammation. Limitations of the present study are the small cohort size showing a broad spectrum of COVID-19 severity and that we analysed a single plasma sample from each patient at individually different phases of their hospitalization. Therefore, a generalization of the results to all patients suffering from COVID-19 is not possible and we cannot discuss data on dynamic alterations of cfDNA over time.

The results provide evidence that cfDNA is a liquid biopsy marker potentially predicting disease severity of COVID-19. Our findings are in agreement with recently reported data on the predictive value of circulating (mitochondrial) cfDNA for COVID-19 outcome (20, 21). Significant correlations were further reported between total cfDNA and LDH (preprint medRxiv, https://www.medrxiv.org/content/10.1101/2020.07.27.20163188v1), a biomarker of cell damage, similar to circulating cfDNA. LDH is commonly elevated in severe COVID-19 and is increased in non-survivors (23). As the relative range of cfDNA concentrations determined in COVID-19 patients in our study was broader than the range of LDH, cfDNA may be utilized to discriminate different grades of COVID-19 severity more accurately than LDH. Testing this hypothesis will be the subject of future studies. It should be noted that cfDNA is not only a biomarker for upstream pathophysiological mechanisms but has been also proposed to trigger specific downstream effects. For instance, cfDNA was shown to be an immune system regulator (24) with distinct immunoregulatory properties in healthy and diseased individuals (25). It has also become clear that cfDNA is among the factors contributing to neutrophil extracellular trap (NET) formation. NET plays a key role in immunothrombosis and was demonstrated to be consistently increased in COVID-19 and linked to disease severity (26, 27). Indeed, complications such as acute arterial thromboembolism have been reported in COVID-19 (28) and inflammatory cells are prominent in arterial thromboembolic material from COVID-19 patients (29). However, recent data did not provide evidence for classic thrombotic microangiopathy in COVID-19 (22) which is in agreement with our observation that higher cfDNA concentrations were not associated with thromboembolism. One hypothesis is that laboratory hallmarks of thrombotic microangiopathy are lacking in COVID-19 due to its restriction to the pulmonary microcirculation (22) which distinguishes COVID-19 pulmonary pathology from that of equally severe influenza virus infection (30). It has been hypothesized that neutrophils can amplify pathological damage and aggravate a hyperinflammatory state (31). Moreover, SARS-CoV-2 was found to induce the release of neutrophil extracellular traps (NETs) by neutrophils (32). It remains to be clarified whether specific pathways downstream of cfDNA are crucial for hyperinflammation and COVID-19-associated coagulopathy following SARS-CoV-2 infection. However, current knowledge is compatible with the hypothesis that cfDNA triggers both hyperinflammation and coagulation (Fig. 1).

**Fig. 1.**
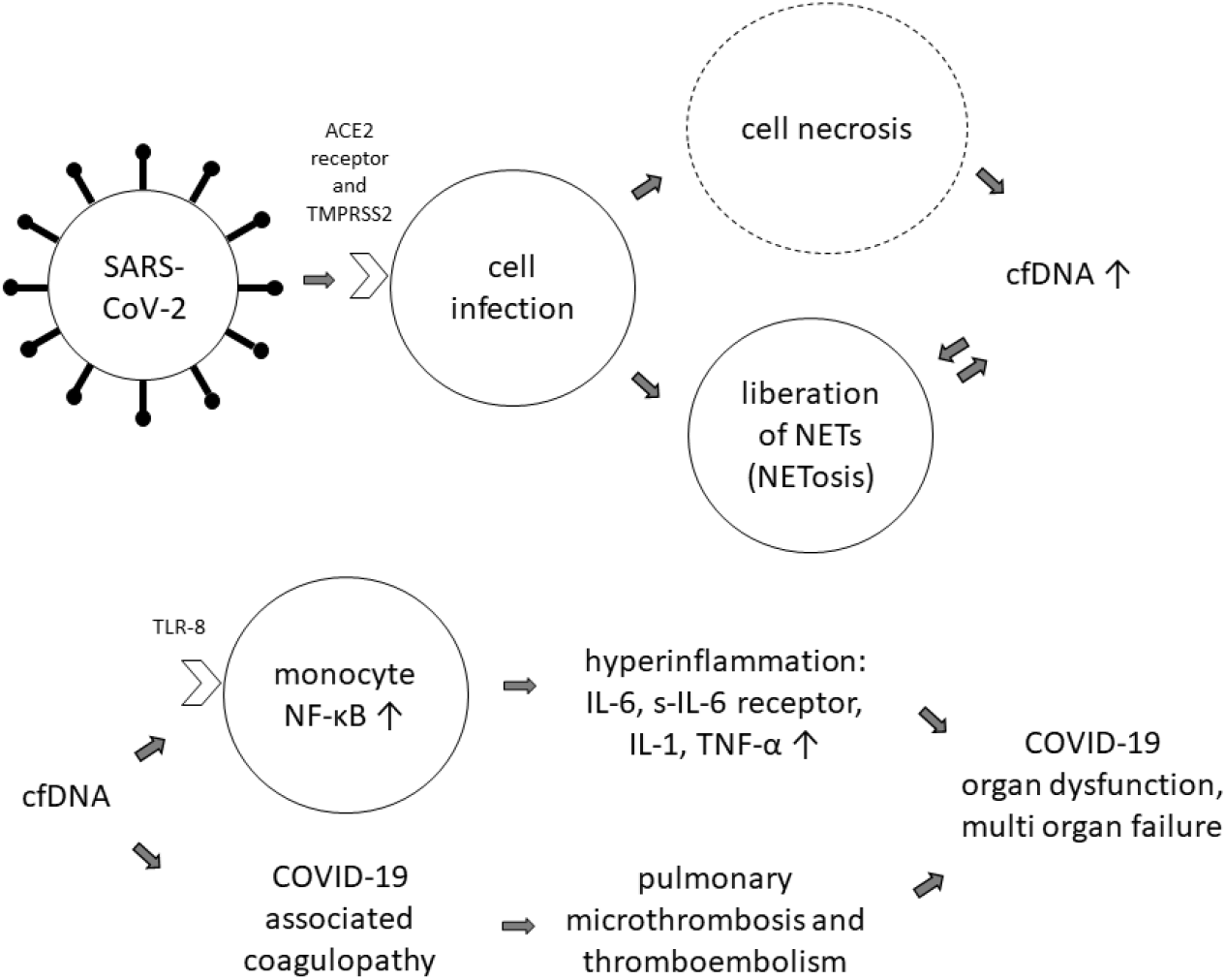
Hypothetical role of cfDNA in COVID-19. Abbreviations: *NETs*, neutrophil extracellular traps; *NF-κB* (Nuclear Factor Kappa-light-chain-enhancer of activated B-cells); *IL-6* (Interleukin 6), *IL-1* (Interleukin 1), *TNF-α* (Tumor Necrosis Factor Alpha)

## Conclusion

cfDNA concentration in blood plasma samples from COVID-19 patients correlated with the occurrence of clinical complications and disease severity. Clinical laboratory measurements for D-Dimer and CRP showed the highest correlations with cfDNA levels. Our data allow to generate hypotheses for prospective trials to confirm the predictive value of cfDNA in COVID-19 when samples are taken at defined time points of the disease course.

## Data Availability

data are available from the corresponding author upon reasonable request

## Notes

### Competing Interest Statement

The authors have declared no competing interest.

### Funding Statement

not applicable

### Author Declarations

The study was approved by German law [Landeskrankenhausgesetz] in accordance with the Declaration of Helsinki and by the local Ethics Committee of Landesaerztekammer Rheinland-Pfalz (reference numbers 2020-15116-retrospective).

